# Required COVID-19 Vaccination Certification to Attend Certain U.S. Professional Football Games in Fall 2021: A Natural Experiment

**DOI:** 10.1101/2022.02.13.22270900

**Authors:** Leon S. Robertson

## Abstract

Four U.S. National Football League teams required vaccination certification against the SARS-CoV-2 virus of attendees at home games during the 2021 season. Using daily data on confirmed cases and vaccinations in counties surrounding these stadiums and stadiums that did not require certification, this study estimates the effects of the certification policy. Ordinary least squares regression was used to estimate the change in community spread of the virus after home games and away games relative to weeks that the teams did not play (bye weeks).

Compared to counties in metropolitan areas near stadiums with no certification requirement, counties near stadiums that had a vaccination requirement had significantly less cases 14 days after home games. In the six weeks leading up to the beginning of the season, percent vaccinated increased in counties that were near stadiums requiring vaccination certification only if the prevalent preseason vaccination rate was relatively low. Required vaccination certification at venues for large gatherings appear to slow virus spread generally in nearby communities and increases vaccination percentages in areas with lower prevalent vaccination percentages.

## Introduction

Professional football fans in the U.S. were allowed to attend games during the 2021 season after the 2020 games were played with limited numbers of fans allowed to enter the stadiums to reduce spread of the pandemic virus SARS-CoV-2. One study of counties with limited in-person attendance at professional and NCAA college games in 2020 compared to matched counties with no games found no increased spread in cases after games^1^. A second study that included all counties in metropolitan areas where National Football League (NFL) games were played reported more cases associated with games in 2020^2^.

NFL teams allowed full attendance in the fall of 2021, averaging more than 67,000 people per game ^3^. Television displays of attendees during games suggested that the vast majority of attendees were not wearing masks. Many fans at games could be seen screaming for the home team and, if infected with the SARS-CoV-2 virus, potentially spreading it to the people in front of them. The newly infected could then take the virus home to their families, neighbors and friends.

To reduce the risk of spreading the virus, the NFL urged potential attendees to stay at home if they were exposed to COVID-19 within 14 days before a game or had a variety of symptoms within the past 48 hours. They were also asked to stay at home if they were not vaccinated or had a positive test ^4^. Four teams (Buffalo, Las Vegas, New Orleans and Seattle) required documentation of vaccination upon entry to their stadiums^5^. During October 10 through December 14, 2021 each team had one “bye week”, that is, no game^6^.

These conditions created a natural experiment. Did the virus spread more or less in metropolitan areas surrounding a given team’s stadium after home games and away games relative to weeks with no games? During away games, people often gather in homes and bars to watch the games on television so a clear hypothesis regarding the potential relative effect of home versus away games is problematic. Since vaccination reduces the risk of infection more than 90 percent^7^, counties in metropolitan areas of the teams requiring vaccine certification should experience less COVID-19 cases two weeks after home games than areas where the home team does not require game attendees to be vaccinated. This study examines change in reported COVID-19 cases in metropolitan counties after two weeks following home and away games of their local professional football teams. Preseason changes in vaccination percentages in metropolitan areas with and without the vaccine certification requirement were also examined.

## Materials and Methods

Daily cumulated COVID-19 confirmed cases in U.S. counties were downloaded from USAfacts.org ^8^. To avoid spikes related to daily reporting variation, seven-day moving averages of new cases per day were calculated for each county. Counties within metropolitan statistical areas^9^ that included NFL teams were identified and matched to the daily case data for those counties. The Baltimore-Washington, Los Angeles and New York areas were excluded because each has two teams within their metropolitan statistical areas. The dates of home games, away games and bye weeks of the remaining teams^10^ were matched to the to the daily case data. The number of cases on game day and Sunday of bye weeks of each county in a given metropolitan area and the number 14 days later were included in the data set.

Ordinary least squares regression was used to estimate the cases in 14 days after a home game with vaccine restrictions, a home game without vaccine restrictions, and an away game, each relative to bye weeks. Logarithms of cases were used to reduce skewed distributions. The form of the equation is:

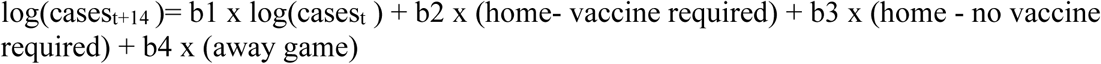

Where t = game days and the home and away variables are 1 if applicable, otherwise zero.

Inclusion of cases on game day corrects for the status of infections in the county at the time of the game. The analysis only included games during the period that included bye weeks, October 18, 2021 – December 11, 2021. Since inclusion of a variable for bye weeks would over-specify the equation, the results are an estimate of the influence of attendance conditions on virus spread 14 days after game days relative to no game, home or away, for a given team.

To examine the changes in vaccination rates in the subject counties prior to the first game, vaccination rates on the regular season opening day were compared to rates the first of August prior to the season. The data were downloaded from the U.S. Centers for Disease Control and Prevention.^11^

## Results

The regression coefficients are displayed in Table 1. Cases in counties after home games in areas near stadiums that had no vaccine certification requirement were a bit lower than expected from cases during bye weeks but the confidence intervals overlap zero. In areas near stadiums that had a vaccination certification requirement, cases were significantly less after home games than after away games and bye weeks. Generally, irrespective of vaccine requirements, cases were somewhat lower after away games than two weeks after a bye.

**Table 1.**
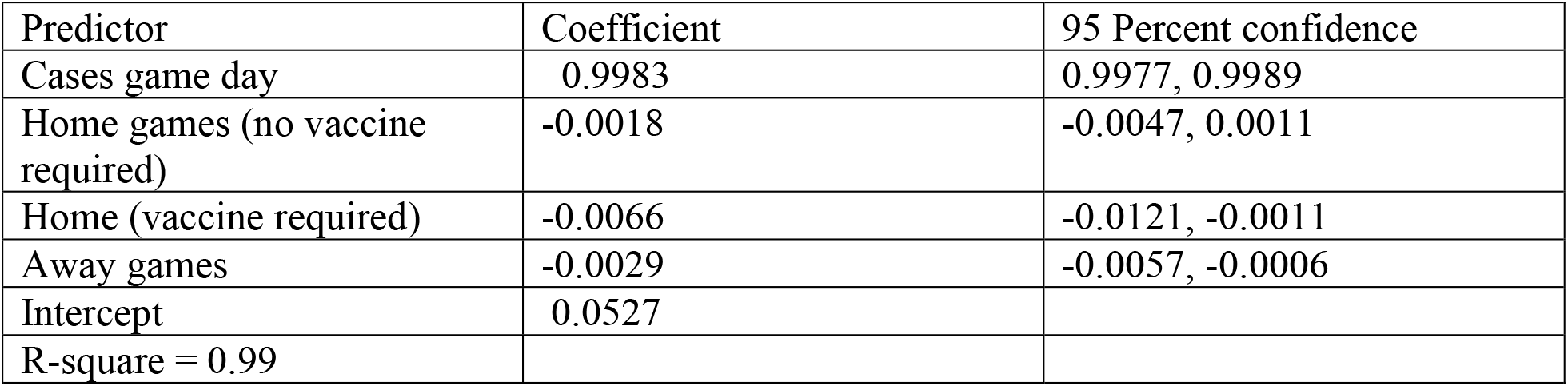
Regression coefficients predictive of COVID-19 New Cases 14 days after NFL games in metropolitan areas that have only one team – October 18, 2021 – December 11, 2021

| Predictor | Coefficient | 95 Percent confidence |
| --- | --- | --- |
| Cases game day | 0.9983 | 0.9977, 0.9989 |
| Home games (no vaccine required) | -0.0018 | -0.0047, 0.0011 |
| Home (vaccine required) | -0.0066 | -0.0121, -0.0011 |
| Away games | -0.0029 | -0.0057, -0.0006 |
| Intercept | 0.0527 |  |
| R-square = 0.99 |  |  |

Comparisons of the percent vaccinated the first of August and the first regular season game day, September 12, 2021, are presented in Table 2. New Orleans and Las Vegas had lower vaccination rates than Seattle and Buffalo and increased more by the first game than cities without the requirement. Seattle’s increase was near that of those not requiring vaccines and Buffalo’s was less.

**Table 2.** Average percent of persons 18 years and older vaccinated against COVID-19 by 8/1/12021 and at the beginning of the NFL regular season 9/12/2021 among counties in metropolitan areas where teams required attendee vaccination and others.

| Date | New Orleans | Las Vegas | Seattle | Buffalo | Other |
| --- | --- | --- | --- | --- | --- |
| 9/12/2021 | 55.1 | 50.6 | 66.9 | 64.4 | 44.8 |
| 8/1/2021 | 48.3 | 46.2 | 63.1 | 61.8 | 41.2 |
| Difference | 6.8 | 4.4 | 3.8 | 2.6 | 3.6 |

## Discussion

These results suggest that NFL warnings to self - screen for COVID-19 infection risk before attending games had little, if any, effect on subsequent spread of the virus as indicated by the lack of significant change in viral transmission after home games in stadiums without a vaccination requirement. When teams required evidence of vaccination to enter their stadiums and played at home, the subsequent two-week spread of the virus was substantially lower. This finding suggests that fans were at less risk at a venue where everyone except those using forged documents was vaccinated rather than pursuing whatever activities were prevalent on Sundays during bye weeks. Apparently, gatherings to watch away games involves less risk of spreading infections than activities during bye weeks.

The results regarding changes in vaccination rates indicate that metropolitan areas with lower pre-requirement vaccination rates benefited more from the requirement. Since the areas where teams have no requirement have even lower vaccination rates on average, a requirement in those stadiums would likely be beneficial to their communities. Vaccination requirements to enter other venues that attract large crowds would likely be beneficial as well.

## Data Availability

All data produced in the present study are available upon reasonable request to the authors.

